# High-dose glucocorticoids, pain, and adverse events. Protocol for an analysis of a natural experiment of patients undergoing hip and knee arthroplasty

**DOI:** 10.1101/2025.11.11.25339982

**Authors:** Jens Laigaard, Marcus Ølgaard Møller, Søren Overgaard, Ole Mathiesen, Anders Peder Højer Karlsen

## Abstract

**Introduction:** High-dose glucocorticoids are increasingly used in patients undergoing primary total hip arthroplasty (THA), unicompartmental knee arthroplasty (UKA), and total knee arthroplasty (TKA). However, the evidence supporting the use of high-dose glucocorticoids to manage acute postoperative pain is based mainly on randomised controlled trials, which may have limited external clinical validity due to strict exclusion criteria. In Denmark, the treatment was implemented stepwise, comprising a natural experiment.

**Objective:** To examine the real-world effect of a single dose of high-dose glucocorticoids on postoperative opioid consumption and adverse events in patients undergoing THA, UKA, and TKA.

**Methods:** This protocol describes a natural experiment study that will be reported as a target trial emulation, i.e. by attempting to mimic a randomised clinical trial.

*Inclusion and exclusion criteria:* All adults (>18 years) undergoing primary, elective THA, UKA, and TKA in the Capital Region and Region Zealand in Denmark are eligible

*Intervention:* The ***intended use*** of a single high dose of glucocorticoids, administered after induction of anaesthesia, as part of the standard treatment.

*Comparator:* No administration of glucocorticoids. Patients in the control arm may have received a lower dose of glucocorticoids for postsurgical nausea and vomiting

*Assignment of intervention:* For each hospital, patients who underwent surgery ***before*** the implementation of glucocorticoids serve as controls, while patients operated on ***after*** the implementation constitute the treatment arm.

*Outcomes:* The primary outcome is opioid consumption, expressed in morphine equivalent doses (MEQs) in the first 24 hours after surgery. Secondary outcomes are two *adverse effect-outcomes:* incidence of opioid-related adverse events and days alive and out of hospital at 30 days, and three *efficacy-outcomes:* length of postoperative observation area-stay, length of hospital stay, and worst 0-24-hour pain intensity score measured.

*Data collection:* This study uses data from the TRIPLE-A project (www.triplea.dk), comprising validated electronic health record data.

*Statistical analyses:* The analyses will be based on a pre-defined mixed effects model with hospital as a random effect and adjusted for important presurgical (and thus pre-intervention) variables. A per-protocol population will be analysed as a sensitivity analysis. For the primary outcome, a difference of 5 MEQs between treatment arms is considered clinically important.

**Knowledge dissemination:** The results will be shared at conferences and made publicly available.

**Registration:** *https://doi.org/10.1101/2025.11.11.25339982*

## Introduction

Total hip arthroplasty (THA), unicompartmental knee arthroplasty (UKA), and total knee arthroplasty (TKA) are common orthopaedic procedures in patients with osteoarthritis. Postoperative multimodal pain management is important for patient recovery and satisfaction. This includes high-dose glucocorticoids, which have been investigated for their ability to reduce inflammation and pain. After two randomised controlled trials (RCTs) showed benefit,^1,2^ high-dose glucocorticoids have been implemented as routine treatment for THA, UKA, and TKA in Denmark.^3^ However, due to concerns about adverse effects, e.g. increase risk of infection, the treatment was only step-wise implemented, as more evidence emerged.^4–7^ Still, the efficacy and adverse event profile is primarily based on data from RCTs, often with strict exclusion criteria and missing data,^8,9^ rather than ‘real-world’ evidence.^10^

In a parallel project,^11^ we have mapped the time of implementation of high-dose glucocorticoids at Danish orthopaedic departments to evaluate if the treatment reduced long-term opioid use and affected the number of days alive and out of hospital. For that project, we are unable to determine if the patients did in fact receive the treatment. Moreover, we did not plan examine short-term outcomes because we were unable to include in-patient data from the electronic health record systems. Here, using data from the AI and Automation in Anaesthesia (TRIPLE-A) database, we can also investigate the effect of high-dose glucocorticoids on important short-term outcomes.^12^

We aim to examine the real-world effect of high-dose glucocorticoids on postoperative opioid consumption and adverse events in patients undergoing primary THA, UKA, and TKA.

## Methods

This protocol describes the methods for a natural experiment study. The study will be reported following the target trial emulation framework.^10,13^ The study was listed at the Capital region of Denmark’s regional research listing (identifier p-2025-20115), the Ethical Committee of the Capital Region of Denmark waived ethical approval for this study (identifier F-25000312, 2 January 2025), and we were granted access to individual patient files approval from the Capitol Region Team for Medical Records Data (identifier R-25000339, 20 March 2025, with update for the specific subproject on 21 August 2025).

### Study setting

The patients considered for this study underwent primary THA, UKA, or TKA surgery at public hospitals in the Capital Region and Region Zealand in Denmark from 2017 to 2025. All residents in Denmark have equal access to free-of-charge healthcare financed by general taxes. Each year, around 10.000 patients undergo primary THA, UKA or TKA at public hospitals in the Capital Region and Region Zealand.^14,15^ Enhanced Recovery After Surgery (ERAS) guidelines are widely implemented and half of the patients are discharged home the day after surgery. ^14–16^

### Inclusion and exclusion criteria

All adults (>18 years) patients undergoing primary, elective THA, UKA, and TKA in the Capital Region and Region Zealand in Denmark are eligible

We will adjust for calendar time, as well as important co-interventions, to account for possible time bias. Moreover, because we believe high-dose glucocorticoids, being a simple pharmacological intervention, were relatively abruptly implemented, we will assume that there is no need for a ‘training’ or washout period.

### Intervention

The intervention the ***intended use*** of a single high dose of glucocorticoids, administered after induction of anaesthesia, as part of the standard treatment. A high-dose steroid dose was defined as IV/oral administration of ***at least*** 0.2 mg/kg or 16 mg dexamethasone/dexamethasone phosphate, 80 mg methylprednisolone, or 105 mg prednisolone (https://www.mdcalc.com/calc/2040/steroid-conversion-calculator).

The intervention was given along with other analgesic interventions that were routinely given to THA and TKA patients in Denmark during the study period, including paracetamol, NSAIDs, gabapentin, opioids, and, for TKA patients, local infiltration analgesia.

### Comparator

The comparator is no administration of glucocorticoids during anaesthesia. Patients in the control arm may have received a lower dose of glucocorticoids for postsurgical nausea and vomiting. Typical anti-emetic doses are 4-8 mg dexamethasone.^17^

### Data sources

For this project, we will use data available from the TRIPLE-A database.^12^ The TRIPLE-A database include hundreds of variables on well over one million surgical procedures performed in the Capital Region and Region Zealand in Denmark from 2017 to 2025, extracted from the electronic health record system (EHR) Sundhedsplatformen (Epic Systems Corporation, WI, USA).

### Assignment of intervention

For each hospital, patients who underwent surgery ***before*** implementation of glucocorticoids serve as controls, while patients operated ***after*** implementation constitute the treatment arm, regardless of their actual treatment.^18,19^ Thus, treatment allocation is non-random and open label. Because certain patients (for example, those with insulin-dependent diabetes) may have been excluded from receiving glucocorticoids according to local protocols, we will report the proportion of patients in the treatment arm who did not receive the treatment (i.e. “protocol violations”).

### Outcomes

#### Primary outcome

Opioid consumption, expressed in morphine equivalent doses (MEQs) in the first 24 hours after surgery. We included opioid doses administered pre-emptively at the end of surgery, adjusting for time interval between administration and the actual end of surgery using the drug-specific half-life.^12^ We used the opioid conversion table that is implemented in the TRIPLE-A platform for conversions to MEQs.

#### Secondary outcomes

We chose five secondary outcomes:

- Worst postoperative pain intensity during 0-24 hours after surgery, measured with the 0-10 numerical rating scale (NRS).
- Proportion of patients with at least one opioid-related adverse event (ORADE) during 0-24 hours after surgery.
- Length of stay in the post-anaesthesia care unit (PACU), measured in minutes.
- Length of hospital stay, measured in hours from end of surgery to hospital discharge.
- Days alive and out of hospital at 30 days

### Data collection

Please see the TRIPLE-A project website (currently www.cepra.nu/triple-a, until www.triplea.dk is launched in January 2026) for how each variable is assessed.^12^ In brief, the data quality is dependent on the registrations in the electronic health record system. Administration of medications, surgeries, admissions, readmissions, and length of stay-variables are generally of very high quality, while health-care worker-dependent variables, such as pain scores, can be subject to errors and bias. For this project, we manually validated the treatment variable (administration of high-dose glucocorticoids) and important co-intervention variables (administration of paracetamol, NSAID, opioids) in 50 randomly selected patients without finding any errors.

### Sample size

The sample size is based on convenience rather than power and the confidence intervals (CIs) will reflect the uncertainty of the estimate. However, using the CRTsize package in R,^20^ we anticipate a power of 95% (5% false-negative rate) for detection of a 5 MEQ difference, assuming a standard deviation of 10 MEQ, 14 paired clusters (7 sites) with 50 patients, an intraclass coefficient of 0.1, and a 5% false-positive rate.

### Statistical analysis plan

The data will be stored and analysed at a logged encrypted drive with the statistical software R (R Core Team, Vienna, Austria).^21^ The dataset will be deleted after publication of the manuscript.

#### Missing data

We will check for the proportion of missing outcome data before conducting analysis. The analysis for a given outcome will not be conducted if the missingness is unbalanced. If the missingness is balanced between treatment arms, we will impute missing data using multiple imputation with the mice package.^22^ The number of patients contributing data to each analysis will be reported for each analysis.

#### Modelling

The analyses will be based on a pre-defined mixed-effects model using the lme4 package^23^, adjusted for important presurgical (and thus pre-intervention) variables. For continuous outcomes:

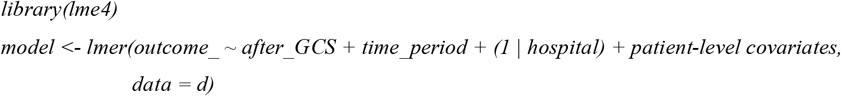

Where **outcome_** is the continuous outcome of interest, **after_GCS** is the DiD variable indicator of whether high-dose glucocorticoids were implemented at the hospital where the patient underwent surgery, **time_period** is a categorical (factor) variable with a level for each interval between time of implementation, **hospital** is a categorical (factor) variable indicating which hospital the patient underwent surgery at, and **patient-level covariates** are the following variables, including as fixed effects: Age (continuous variable), Sex (binary variable), Type of surgery (binary variable: THA/TKA), Type of Anaesthesia (binary variable: General/Spinal), Paracetamol, NSAID and/or gabapentin as premedication or given intraoperatively (continuous variable with 0-3 medications), Local infiltration analgesia (binary variable), Preoperative chronic opioid use (binary variable) and Number of psychiatric diagnoses on admission (continuous variable). Continuous outcomes are reported as mean difference with 95% (primary outcomes) or 99% (secondary outcomes) confidence intervals (CIs).

Similarly, binary outcomes are reported as absolute risk difference and the relative risk with 99% confidence intervals. First, a generalized linear mixed model with a logistic link function is fitted:

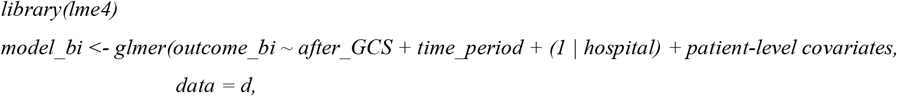

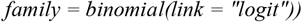

The average treatment effects and confidence intervals are estimated with the *marginaleffects* package^24^:

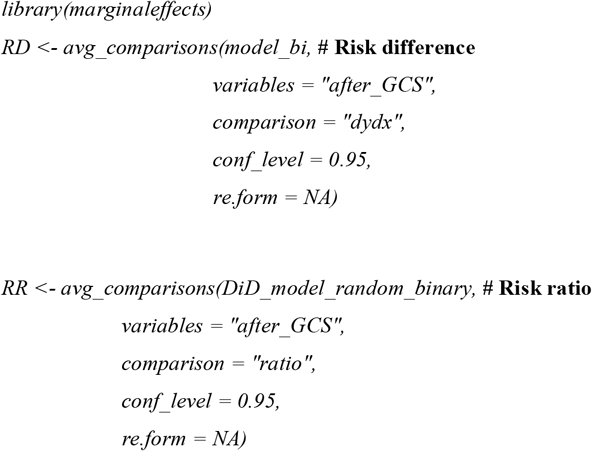

#### Thresholds for significance

The minimal important difference is considered a 5 MEQ absolute difference in opioid consumption within the first 24 hours after surgery.^25^ For the primary outcome, a P-value below 0.05 is considered statistically significant and the outcome will be reported with 95% confidence interval. For secondary analyses, we will apply Bonferroni correction to manage the risk of false-positive, i.e P-values below 0.05/5 = 0.01 are considered statistically significant. Secondary outcomes are reported with 99% confidence intervals.

#### Sensitivity analyses

We will conduct per-protocol sensitivity analyses, excluding patients who have not received the intervention after implementation and patients who have reived the intervention before the implementation. For example, patients with diabetes may not have received high-dose glucocorticoids immediately after implementation due to concerns over spikes in blood glucose

#### Planned Tables and Figures

We will present a table of demographic information, intraoperative, and perioperative details for both groups, including – but not limited to – age, sex, ASA-scores, BMI, type of surgery, type anaesthesia, propofol and remifentanil infusion rates, and the proportion of patients treated with high-dose glucocorticoids, paracetamol, NSAID and LIA.

To visualise and examine the findings and underlying trends, we will use statistical process control in control charts with monthly data relative to the time of implementation. We will chart all assessed outcomes, the proportion of patients who are administered high-dose glucocorticoid, and the proportion of patients who are administered important co-interventions, such as pre- or intraoperative paracetamol, NSAIDs, LIA, and spinal anaesthesia. We will apply the Anhoej rules (‘unusually’ long runs and ‘unusually’ few crossings of the median) and Shewhart’s 3-sigma rule (values outside +/-3 SDs) to assess for non-random patterns in the distribution of data points. For continuous measures, we will plot the monthly mean (Xbar-chart). For binary measures, we will plot the monthly proportion of patients.

#### Data sharing

We refer to the TRIPLE-A project group regarding data availability.^12^ The statistical code can be obtained from the corresponding author upon reasonable request.

#### Knowledge dissemination

The results will be shared at conferences and made publicly available either through open-access publication or through publication at a preprint server, e.g. MedRvix.org.

## Data Availability

All data produced in the present work are contained in the manuscript.

## Roles

Jens Laigaard and Anders Peder Højer Karlsen drafted the protocol with inputs from Ole Mathiesen and Søren Overgaard. Jens Laigaard will handle the data, conduct and interpret analyses, supervised by Anders Peder Højer Karlsen. Jens Laigaard will draft the manuscript, which will be critically revised by all authors.

